# Multi-species plasmids and *K. pneumoniae* clonal spread driving *bla*_NDM_ outbreak across seven UK healthcare sites

**DOI:** 10.1101/2025.05.09.25327284

**Authors:** Caitlin Duggan, Charlotte Brookfield, David Lawrie, Victoria Owen, Timothy Neal, James Cruise, Alice J Fraser, Lewis Kelly, Fabrice E Graf, Daire Cantillon, Joseph M Lewis, Thomas Edwards, Eva Heinz

## Abstract

**Background:** The NDM carbapenemase enables Gram-negative bacteria to hydrolyse almost all β-lactam antibiotics. In 2023, a large increase of *bla*_NDM_ in clinical isolates was identified in a clinical microbiology laboratory serving hospitals and GP practices in Merseyside, UK, raising concerns of a larger outbreak.

**Methods:** We used short read whole-genome sequencing and bioinformatic analyses to identify the outbreak(s) and identify potential links by combining with anonymised, routinely collected clinical metadata. A subset of isolates were long read sequenced to analyse resistance genes and plasmids at maximum resolution.

**Findings:** The outbreak included Enterobacter hormaechei, Klebsiella pneumoniae, Escherichia coli, Citrobacter freundii, Citrobacter brakii, and Enterobacter kobei and 28 sequence types. K. pneumoniae ST101 represented the only major clonal expansion (18/21 K. pneumoniae isolates), and combination with metadata strongly suggested a multisite hospital-associated outbreak. We identified four variants of bla_NDM_, and long read sequencing indicated bla_NDM-1_ spread across species was driven by an IncHI2/IncHI2A plasmid, representing 82% of bla_NDM1_ isolates.

**Interpretation:** This study captures an outbreak of *bla*_NDM_ positive *Enterobacterales* across eight healthcare settings and highlights the complex epidemiology of spread mediated by both expansion of successful lineages (*K. pneumoniae* ST101) and multi-species plasmids (IncHI2/IncHI2A). It also highlights the challenge recognising and tracing an outbreak linked to patients moving across different healthcare systems and the importance of linked surveillance systems.

**Funding:** We acknowledge funding from Wellcome (217303/Z/19/Z, to EH), BBSRC (BB/V011278/1, BB/V011278/2, to EH) and an Academy of Medical Sciences Springboard award (REF: SBF009\1181, to TE).

**Research in context:** *Evidence before the study:* We searched the pubmed database for similar reports using the search terms ((NDM) OR (New-Delhi metallo-β-lactamase)) AND (Enterobacterales) AND (hospitals) AND (outbreak) AND (plasmid) AND ((WGS) OR (whole genome sequencing)) (date of search 08. 03. 2026). We identified 75 studies, from 2019 to 2026, mainly focused on either country-wide surveillance data (n=25), single-species analyses (n=15), or single-hospital outbreaks (n=34). The studies, in particular high-resolution single-hospital outbreak studies, showed the complexity of tracking a plasmid-driven resistance outbreak across multiple species. Country-wide data gave important insights into the ease of spread of Enterobacteriaceae, and the importance of environmental sampling to track transmission, though limited resolution on patient movements.

*Added value of this study:* Our study combines high-resolution patient movement data, whole-genome sequence (WGS) information to track both plasmids and species, with the insight gained via inclusion of several different hospitals, some of which based in distinct administrative systems. Our study adds an overview of the full span of complexity in tracking an outbreak in modern healthcare systems by patients moving between different hospitals and within hospitals between wards on one side, with the resolution necessary to track the Russian doll-like movement of antimicrobial resistance determinants covering movement of insertion elements within plasmids and between plasmids and chromosomes, movements of plasmids between species, and transmission of plasmid-harbouring bacteria between patients, linking back to the clinical complexity.

*Implications of all the available evidence:* Current outbreak investigations usually focus on a specific species with a given resistance profile. Our work shows the relevance to consider mobile resistances in a different framework, i.e. that the movement of the resistance determinant itself needs to be monitored in addition to the putative clonal lineage which is the approach for typical pathogen spread. In addition, we highlight the complexity imposed by a modern clinical healthcare system, where patients need to be tracked across administrative boundaries to ensure capturing frequent transmission routes and identify high-risk points of transmission which might then spread into different hospitals and wards, losing the signal of clustered cases.

**Data sharing statement:** All metadata used in this study is provided in the supplementary tables, and all raw sequence data is available on SRA/ENA under BioProject PRJEB89345; detailed read accessions are given in Supplementary Table 1.

## Introduction

In 2019, an estimated 1.27 million global deaths were attributed to bacterial antimicrobial resistance (AMR), with the highest number of deaths linked to *Escherichia coli* and *Klebsiella pneumoniae* (1). These bacteria, members of the *Enterobacterales* order, are particularly difficult to treat when they produce carbapenemase enzymes which inactivate most beta lactam antimicrobials. They are associated with poor outcomes (2), and have been identified by the world health organisation as critical priority pathogens (3). In England, the incidence of carbapenemase-producing organisms is increasing rapidly, with 7,438 cases of acquired carbapenemases reported to the UK Health Security Agency (UKHSA) in 2024, compared to 666 in 2021 (4). An understanding of the reasons for this rise, and strategies to interrupt transmission, are urgently needed.

Treatment options are particularly limited for metallo-beta-lactamase (MBL) carbapenemase enzymes (5), of which the NDM enzyme is most common in the UK (4), and worldwide (6). Originally described in 2009, the *bla*_NDM_ gene has by now spread across a large range of species with sequence data reporting up to 59 bacterial taxa (7), disseminated on plasmids often with a diverse bacterial host range. Transmission within hospitals is thought to be a key driver of dissemination to a vulnerable host population. Outbreak tracing of CPEs (including NDM producers) has historically focussed on bacterial strain typing to infer transmission between patients or environmental sources (8), but newer sequencing techniques and advancements in typing of mobile genetic elements (MGEs) like plasmids has allowed an assessment of the role of MGEs in driving multispecies spread of resistance genes (9).

In this context, between 2022 and 2023, we noted a year-on-year increase in the number of inpatient samples positive for *bla*_NDM_ in the Liverpool Clinical Laboratories, (LCL). We carried out a retrospective genomic-epidemiologic investigation of *bla*_NDM_-encoding Enterobacteriales to understand the drivers of this increase and the mechanisms of spread to guide prevention efforts.

## Methods

### Study setting

The LCL serves seven adult hospitals (referred to here as hospitals 1-6, and 8) and all GP practices throughout the city. These hospitals include two general acute hospitals with emergency departments and a number of specialty hospitals (Supplementary Table 2), with complex care journeys for patients who may be transferred between sites to access specialty care, and are managed by several NHS trusts.

During the study period, high-risk hospital inpatients were screened for CPE colonisation following UK guidelines (10) which includes rectal swabs, and as appropriate urine or wound samples (hereafter referred to as “screening samples”); clinical samples were also sent from hospital and GP sites when infection was suspected (hereafter referred to as “clinical samples”) which were processed according to standard methods. All meropenem resistant isolates were tested for carbapenemase genes using the Xpert Carba-R assay, combined with chromogenic agar for critical care screens and meropenem for clinical samples, respectively (see supplementary methods).

### Sample and metadata collection

We retrospectively identified all isolates in which *bla*_NDM_ was detected in the clinical laboratory from January 2023 to December 2023 using the laboratory information management system. There were 68 in total, and 63/68 of could be retrieved for further investigation, and passed sample QC including 23 *Enterobacter* spp., 21 *K. pneumoniae,* 15 *E. coli* and four *Citrobacter* spp. Following HRA approval (IRAS ID 345990), we extracted anonymised linked patient metadata from electronic health records for these 63 isolates, including dates and times of hospital ward moves, though these ward move data were only available for hospitals 1,2,3 and 5, which accounted for 60/63 isolates. We defined a strong epidemiologic link between isolates as those cultured from different participants who had been on the same ward on the same day, and a weak link defined as isolates cultured from samples collected from participants who had been on the same ward at any time (i.e. not necessarily on the same day).

### Whole-genome sequence analyses

All samples underwent short-read sequencing on an Illumina platform, and 24 were also sequenced on the Oxford Nanopore MinION platform using the Native Barcoding Kit 24 V14 (SQK-NBD114.24) (ONT, United Kingdom) and protocol. After QC, either short-read only or hybrid genomes (for the isolates with long-read sequencing available) were assembled, and determinants of interest (antimicrobial resistance genes, capsule types plasmid replicons) were determined. Plasmid assemblies were manually inspected, and hybrid-assembled plasmids and gene cassettes were compared for synteny and conservation (details in supplementary methods).

### Phenotyping

For *K. pneumoniae*, string tests were performed according to a previously published methodology (57); and only two isolates encoded for yersiniabactin and yersiniabactin + aerobactin, and a partial *rmpA* gene, resp (Supplementary Table 3). Plasmid conjugation was attempted using an *E. coli* donor (NDM151) and *E. coli* MG1655 recipient using filter assays, and differential selection was used for colony counts and selection of transconjugants which were confirmed by targeted PCRs (Supplementary Table 4). The minimum inhibitory concentration of meropenem was identified for the donor, recipient and a transconjugant using MIC broth microdilution. The ATCC25922 *E. coli* strain was used as a quality control organism, endpoints were interpreted according to the EUCAST clinical breakpoints (v.15.0).

### Ethics statement

No identifiable data was analysed during this project. As an anonymised use of routinely collected data and following UK Health Research Agency (HRA) guidance the study does not require formal research ethics committee review. It was approved by HRA (IRAS ID 345990).

## Results

### Sample and participant details

Between Jan-Dec 2023, a total of 68 isolates were identified as NDM-producing Enterobacterales in the clinical laboratory, and 63 could be retrieved and passed QC for further analysis. These 63 isolates were collected from 61 participants, with one environmental isolate (Table 1). Most isolates from humans were from colonisation screens (56/63, 89%); six were clinical diagnostic samples (2 urine; 1 tissue; 2 wound swab; 1 cerebrospinal fluid), and one environmental sample derived from an outbreak investigation at hospital 2 (a shower trap). Over half of the isolates were from the two acute hospitals (Hospitals 1 and 5, 49% and 21% respectively) and the specialist oncology hospital (Hospital 2, 17%); one isolate each was derived from Hospitals 4, 6 and 8, and one GP practice.

**Table 1:** Clinical details of sampled population. This included 61 participants, contributing 62 samples, and one environmental sample. *Duration of hospital admission/prior hospitalisation only available for Hospitals 1,2,3,5 representing 58/61 participants.

| Variable | Value |
| --- | --- |
| **Participant Age\* (years)** |  |
| 0-9 | 1/61 (2% [0-9%]) |
| 20-29 | 1/61 (2% [0-9%]) |
| 30-39 | 9/61 (15% [7-26%]) |
| 40-49 | 6/61 (10% [4-20%]) |
| 50-59 | 13/61 (21% [12-34%]) |
| 60-69 | 7/61 (11% [5-22%]) |
| 70-79 | 14/61 (23% [13-35%]) |
| 80-89 | 7/61 (11% [5-22%]) |
| 90-99 | 3/61 (5% [1-14%]) |
| **Location of sample isolation** |  |
| Hospital 1 | 31/63 (49% [36-62%]) |
| Hospital 2 | 11/63 (17% [9-29%]) |
| Hospital 3 | 4/63 (6% [2-15%]) |
| Hospital 4 | 1/63 (2% [0-9%]) |
| Hospital 5 | 13/63 (21% [11-33%]) |
| Hospital 6 | 1/63 (2% [0-9%]) |
| Community GP site 7 | 1/63 (2% [0-9%]) |
| Hospital 8 | 1/63 (2% [0-9%]) |
| **Sample type** |  |
| Clinical | 6/63 (10% [4-20%]) |
| Environmental | 1/63 (2% [0-9%]) |
| Screening | 56/63 (89% [78-95%]) |
| **Other clinical details** |  |
| Duration of hospital admission* (Median [IQR] days) | 23 (10-44) |
| Hospitalisation within prior 6 months* | 36/58 (62% [48-74%]) |
| Previous negative CPE screen | 35/61 (56% [43-69%]) |
| Days since negative CPE screen (Median [IQR]) | 136 (59-228) |

Age and gender data were available for all 61 participants (Table 1), and hospital and ward admission/discharge dates were available for all those admitted to Hospitals 1,2,3 and 5 (58/61). This is a highly healthcare-exposed cohort where hospitalisation in the 6 months prior to CPE isolation was common (36/58, 62% of participants with available data) and median (IQR) duration of hospitalisation for the episode where CPE was isolated was 23 (10-44), compared to an England mean duration of hospital stay of 8.3 days in 2022(11). The participants were admitted to 65 different areas in total, with a median of 3 (range 1-9) per participant. The most common admission was to critical care (H1_KH45), operating theatre (H1_CX45), an acute assessment unit (H1_RX30), and a surgical ward (H1_OV11), all within Hospital 1 (Supplementary Figure 1). 16/58 (28%) of participants were admitted to two or more hospitals (Supplementary Figure 2), with 13 to two and three participants to three hospitals. Even with 58 patients, the network of ward movements within and between the four hospitals with available data formed a complex structure with clusters of highly connected wards (e.g. ICU [H1_KH45], theatres [H1_CX45], theatre recovery [H1_YV45], surgical ward [H1_YV45], Supplementary Figure 3).

Strong epidemiologic links were identified for 24/58 (41%) patients with location data (Figure 1B, D), consistent with possible transmission events, though they included isolates of different species (Figure 1E). These formed two large clusters (8 and 6 isolates, the larger including isolates from multiple hospitals), one smaller cluster (three isolates) and three isolate pairs (Fig 1E). Isolates with strong links were found in all hospitals with > 1 isolate but were concentrated in Hospital 1, in particular ward H1_KH45 (Figure 1C, E), a critical care area where CPE screening is mandated for all patients.

**Figure 1:**
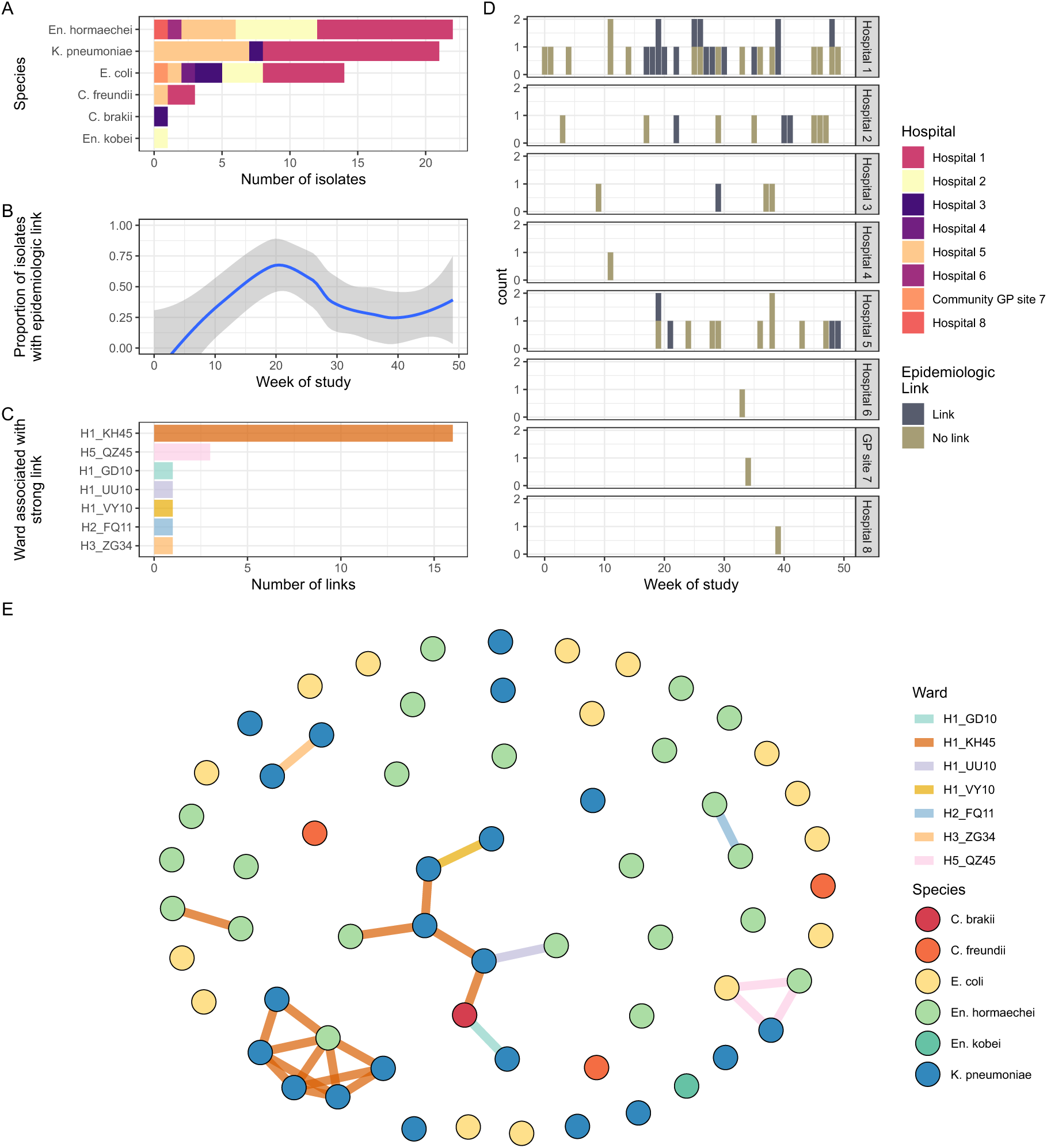
Epidemiological links within and between different species. **(A)** Species distribution of included isolates. **(B)** Proportion of isolates that had an epidemiologic link to at least one other isolate. **(C)** Hospital wards where strong epidemiologic link occurs. **(D)** Isolates cultured per week stratified by site and colour based on at least one vs no epidemiologic link to another sample. **(E)** Network map showing all isolates as nodes (coloured by species) and edges as strong epidemiologic link (coloured by ward).

#### Bacteria: Sequence types and resistance determinants

Having identified possible epidemiologic links, especially including distinct species, we proceeded to WGS and analysis of the 63 isolates to see if the genomic data was consistent with transmission events. This analysis confirmed six species (*En. hormaechei*, n=22; *K. pneumoniae*, n=21; *E. coli*, n=15; *En. kobei*, n=1; *C. freundii*, n=3*; C. brakki*, n=1) and 28 sequence types across all species (Figure 2). The most prevalent ST across the collection was *K. pneumoniae* ST101, representing 86% of all isolated *Klebsiella* in this study (18/21; Fig 2A, E). For the two other prevalent species, *En. hormachei* and *E. coli*, the most prevalent STs only represented 18% and 27%, resp. (4/22 and 4/15). There were only two STs with more than one isolate within the *E. coli.* Across all species, 17 STs were represented by a single isolate; the majority of these (9/17) were *E. coli.* The one participant who contributed two samples was colonised with *E. coli* ST648 on two occasions, two days apart, with 6 core genome SNPs between them. For downstream analysis this was deduplicated by excluding the latter isolate, so all isolates were sampled from different participants.

**Figure 2:**
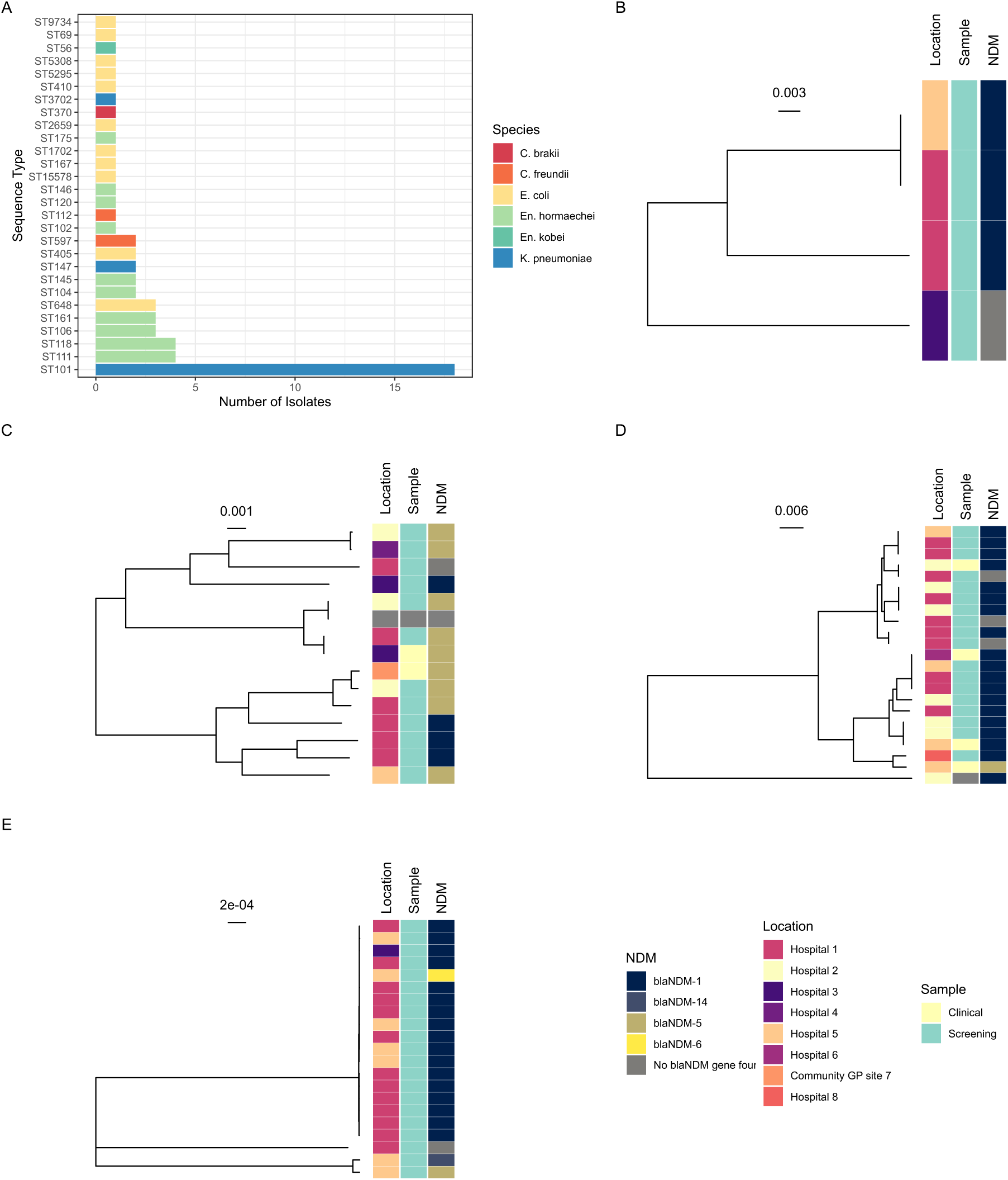
Population structure of the bacterial isolates. **(A)** Sequence types for all included isolates. Phylogenetic trees for **(B)** *Citrobacter* spp, **(C)** *E. coli*, **(D)** *Enterobacter* spp., **(E)** *Klebsiella pneumoniae*. Treescales show substitutions per base for a core gene pseudoalignment; trees are midpoint rooted.

### Multi-hospital clonal transmission driven by *K. pneumoniae*

We then used the WGS data to explore whether the strong epidemiologic links identified could be consistent with transmission, first considering bacteria, then MGE. Of the 24 strong epidemiolgical links we found, 8/24 were of the same species and ST, and all of these involved *K. pneumoniae* ST101.

To assess whether the ST101 *K. pneumoniae* isolates represented a clonal outbreak, we performed a whole-genome SNP analysis, using a high-quality hybrid assembly of the earliest ST101 isolate as reference. Using rPinecone, we identified two sublineages comprised of four and eleven isolates each with fewer than 15 SNPs between isolates, which could be consistent with two putative transmission chains. Three more distantly related ST101 isolates did not belonging to a sublineage (Figure 3A-C). All ST101 *K. pneumoniae* isolates were colonisation screening samples; all sublineage 1 isolates were sampled from Hospital 1 across a five-month period with no strong and only one weak link in H1_KH45, the critical care unit for hospital 1 (Figure 3D-E). Despite a lack of identified epidemiologic links, all four patients carrying sublineage 1 were admitted to hospital in the previous six months: either Hospital 1 alone (two patients), Hospitals 1 and 5 (one patient) or hospital 3 (one patient), possibly consistent with healthcare-associated (rather than community-associated) acquisition.

**Figure 3:**
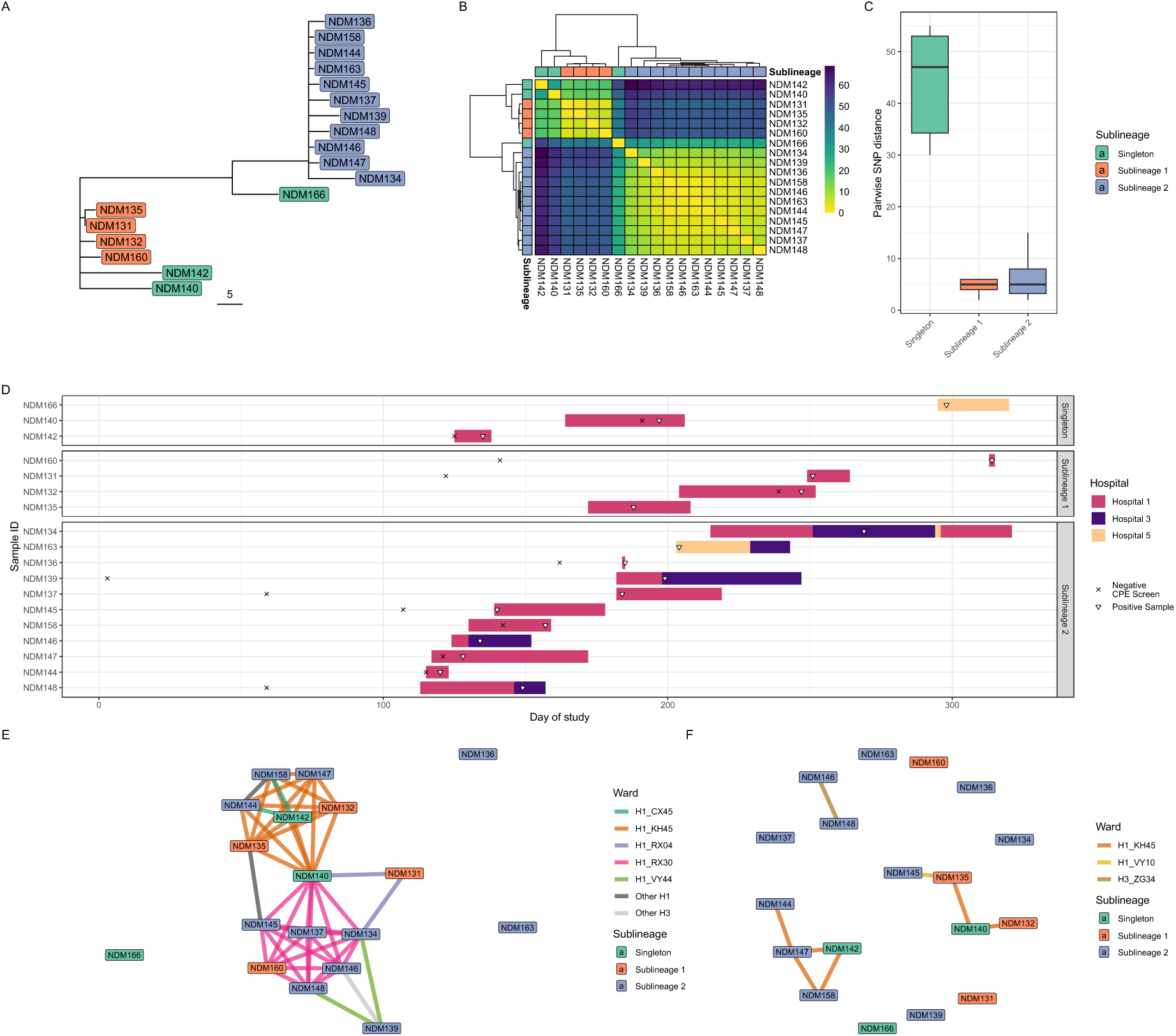
Exploring *K. pneumoniae* ST101 transmission. **(A)** Phylogenetic tree (midpoint rooted) derived from mapping reads to a the earliest ST101 in the study as reference; tree scale here is absolute SNPs; tip colours show rPinecone sublineages. **(B)** Hierarchically clustered SNP-distance heatmap. **(C)** Pairwise SNP distances for the three sublineages. **(D)** Patient admission timeline showing duration of hospital admission for all participants (coloured bars indicating which hospital), time of prior negative CPE screening swab (cross) and positive *K. pneumoniae* ST101 sample (white filled shape). **(E-F)** Weak **(E)** and strong **(F)** epidemiologic links between isolates. Edges represent link (coloured by ward where link took place) and nodes are isolates, coloured by rPinecone lineage.

Sublineage 2 isolates were sampled from three of the hospitals within the study (Hospital 1, 3, and 5), between May and October of 2023. The first two sublineage 2 isolates were identified eight days apart in H1_KH45, the critical care unit for Hospital 1; both patients were negative in earlier screens, on the same ward (Figure 3C), consistent with acquisition on that ward. A third sublineage 2 strain was isolated 29 days after the second from a patient in H1_KH45 who was similarly negative on admission screening, consistent with ongoing person-person transmission or via a shared source. Fourteen days after the second sublineage 2 ST101 isolate was identified at Hospital 1, another sublineage 2 ST101 isolate was identified at Hospital 5 (Figure 3C). No strong epidemiological link was identified, though this participant had been admitted to both Hospital 1 and Hospital 3 within the previous six months. One other strong epidemiological link for sublineage 2 (consistent with person-person transmission) was identified in Hospital 3. Both patients involved were transferred to Hospital 3 from Hospital 1, both had attended Hospital 1 within the previous 6 months, and both were admitted initially to Hospital 1 via ward H1_RX30, an acute medical unit, 11 days apart (representing a weak epidemiological link as per our definition, Figure 2E). A further three sublineage-2 isolates had been isolated from patients who passed through this ward, but not at the same time (i.e weak epidemiologic links as per our definition).

### Co-circulation of highly similar IncHI plasmids encoding for *bla*NDM-1 likely driving multi-species spread

Assessing acquired resistance genes for all WGS data detected 59 genes in total (Supplementary Figure 4A, Supplementary Table 1), with the majority conferring resistance to aminoglycosides and beta-lactams (Supplementary Figure 4A-B). The most common *bla_NDM_* alleles were *bla_NDM-1_* (43/63 isolates) and *bla_NDM-5_* (12/63); and one each for *bla_NDM-6_* and *bla_NDM-14_*. Additional carbapenemase genes of *bla*_IMP-1_, *bla*_VIM-1_, *bla*_VIM-1_ + *bla*_OXA-48-like_, and *bla*_OXA-48-like_ were also observed in *En. hormaechei*, *C. brakii* and *E. coli* in one isolate each (Supplementary Table 1); all these isolates also encoded for a *bla*_NDM._ Whilst the isolates were originally selected for *bla*_NDM_ presence based on hospital diagnostics, no *bla*_NDM_ gene could be detected in WGS data of six isolates, most likely representing loss of the resistance gene or plasmid (Figure 4A).

**Figure 4:**
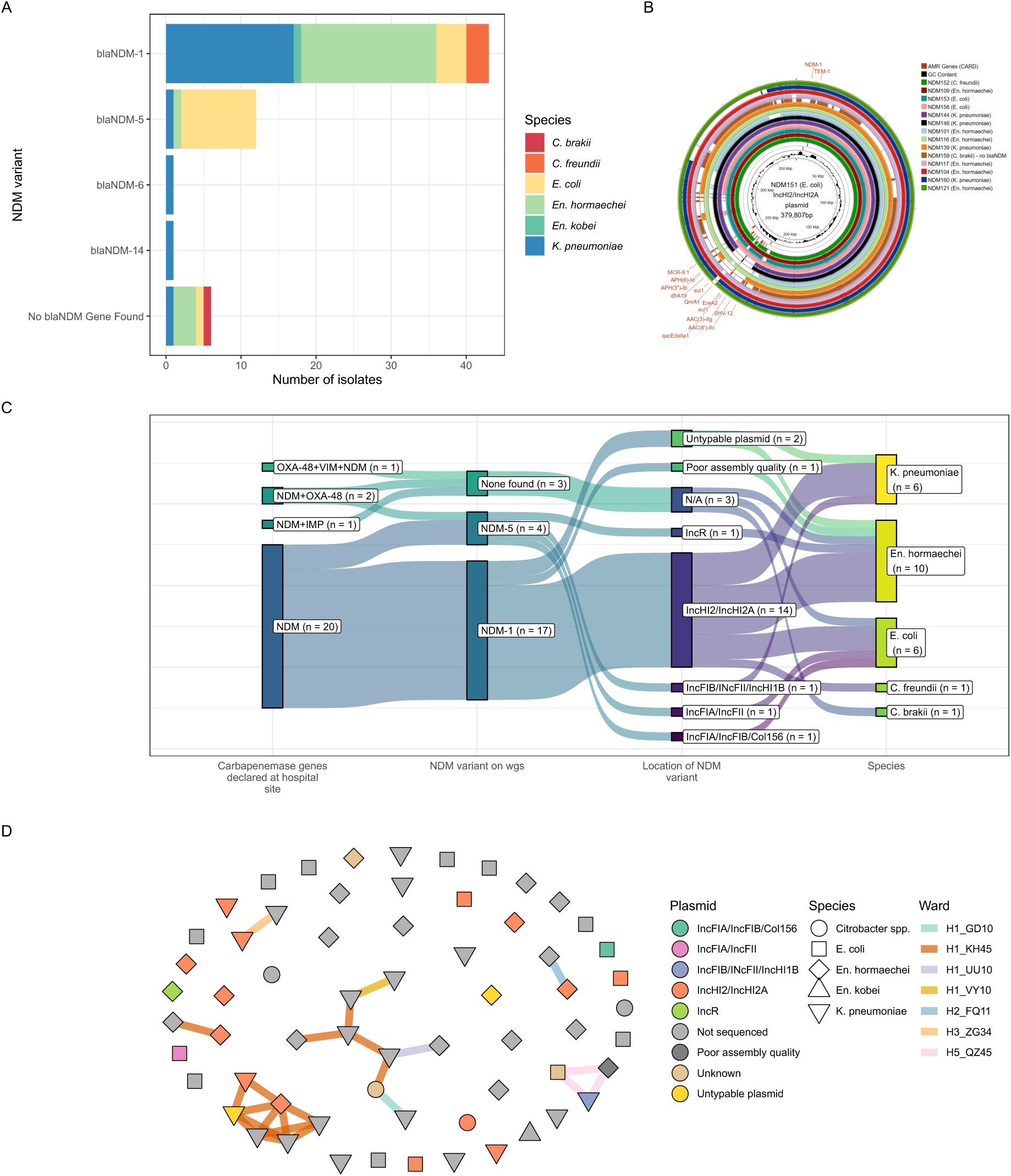
Genomic epidemiology of plasmids identified via hybrid assembly. **(A)** Distibution of *bla_NDM_* alleles in all 63 isolates; **(B)** Proksee visualisation of all 14 IncHI2/HI2A plasmids showing areas of sequence identity > 80% compared to NDM151. **(C)** Sankey diagram showing association of NDM allele, plasmid, and bacterial species. **(D)** Plasmid type (node colour) and bacterial species (node shape) overlaid on the network of strong epidemiological links (edges, coloured by ward of link). In the H1_KH45 ward associated cluster of epidemiologic links (bottom left) two different bacterial species carrying the IncHI2/HI2A plasmid, consistent with a possible transmission event.

To resolve plasmid structures and genetic environments of *bla*_NDM_, we used long-read sequencing of 24 isolates, selected to represent the diversity of the collection in terms of plasmid replicon types (Supplementary Table 1). The majority of isolates with *bla*_NDM-1_ (14/17, 82%) harboured the gene on a IncHI2/IncHI2A plasmid, in a conserved gene cassette flanked either side by IS3000 transposons only 2 isolates encoded this gene on a different (untypeable) plasmid (Supplementary Figure 6), and one had a poor quality assembly so a plasmid could not be identified. The 14 *bla*_NDM-1_ IncHI2/IncHI2A plasmids had a median size of 377Kbp (IQR, 338-385Kbp). In the first outbreak isolate carrying *bla*_NDM-1_ on IncHI2/IncHI2A (*E. coli*), this plasmid additionally harboured multiple other AMR genes, including *bla*_TEM-1,_ *mcr-9* and *bla*_SHV-12_ which we used as reference in a comparison of closed IncHI2/2A sequences from our dataset. (Figure 4B). These had high similarity of plasmid backbone with frequent insertion and deletions.

To explore the possibility that horizontal gene transfer could be responsible for blaNDM transmission events in our hospitals, we looked for strong epidemiological links between isolates with the IncHI2/HI2A plasmid (Figure 4D). Only the 24 isolates subjected to long-read sequencing could contribute to this analysis; of these, 9/24 had a strong link to at least one other isolate. Of these, one isolate pair, including a sublineage 2 ST101 *K. pneumoniae* (NDM144) and a ST145 *En. hormaechei* (NDM145), both carried an IncHI2/HI2A plasmid and had a strong epidemiological link; they were present at the same time on H1_KH45, the critical care unit of Hospital 1.

To test whether the plasmid itself could be driving part of the outbreak, we tested for ability to conjugate and could demonstrate conjugation of the IncHI2/IncHI2A plasmid between an *E. coli* donor (NDM151) to the MG1655 *E. coli* strain recipient and showed that this resulted in meropenem resistant phenotype (Supplementary Figure 7, Supplementary Table 5, 6). Using an *En. hormaechei* (NDM101) and *K. pneumoniae* (NDM160) donor with the MG1655 *E. coli* strain recipient was unsuccessful, likely by lower conjugation rates fore cross-species transfer.

In contrast, *bla*_NDM-5_ was detected on distinct plasmids in each isolate long read sequenced; including IncR, IncFIA/IncFII multi-replicon, IncFIB(pNDM-Mar)/IncFII(29)/IncHI1B(pNDM-MAR) multi-replicon, and a IncFIA/IncFIB/Col156 multi-replicon plasmid (Figure 4C, Supplementary Table 7). The flanking genes of *bla*_NDM-5_ however included a conserved gene cassette of *ble*, *trpF* and *dsbD* that seems to translocate together with *bla*_NDM-5_. (Supplementary Figure 5).

## Discussion

In 2023, an increase of NDM-producing bacteria was observed in the clinical microbiology laboratory serving the majority of people in Merseyside. Clinical investigation revealed that these isolates were largely from hospital inpatients and identified from colonisation screening samples, but were spread across all the inpatient facilities in the city. Routine clinical metadata showed that the patients from whom these samples were collected had complex care journeys with frequent ward and hospital transfers, which took them under the care of different hospital trusts, clinicians, and infection prevention and control teams. At the bacterial level, whole genome sequencing showed that the spreads of the spread of the *bla*_NDM_ gene was driven in part by both clonal spread of *K. pneumoniae* ST101, but also by multispecies spread of a successful IncHI2/IncHI2A plasmid encoding for the *bla*_NDM-1_ gene, in which conjugation was also demonstrated experimentally. There are several implications in these findings.

We identified evidence of patient to patient spread of *K. pneumoniae* ST101 in the critical care unit of Hospital 1 across several months. SNP thresholds to identify likely transmission thresholds of bacteria have been widely debated(12,13), but the ST101 lineage 1 pairwise SNP distances (up to 6 SNPs) are consistent with person-to-person transmission or transmission from a shared source. In recent years, *K. pneumoniae* ST101 have emerged as a potential high-risk CPE clone in European countries, has been strongly associated with spread of CPE(14), carrying blaKPC and blaOXA carbapenemases as well as blaNDM, as in our study. The time scale of persistence of the strain is perhaps suggestive of an environmental reservoir, but we have no systematic environmental sampling to explore this hypothesis. Persistence of *Klebsiella* strains in the hospital environment, e.g. on shared equipment (15) but also particularly in sinks (16), is well described as a cause of outbreaks. The increase in detection of NDMs locally resulted in an infection prevention and control investigation, and a number of interventions applied to reduce transmission (including education and reinforcement of standard IPC measures) after which no more links or *bla*_NDM_ positive isolates were identified within this ward.

In this study*, bla_NDM1_* was very strongly associated with a multireplicon incHI2/IncHI2A plasmid. This was found in multiple bacterial species and in some occasions in patients who shared the same ward at the same time, consistent with transmission events at the level of the bacterial species and/or patient. The fall in price of long-read sequencing technologies makes it feasible to recover full genomic sequences of plasmids at scale and plasmid mediated multispecies CPE outbreaks are increasingly recognised (17), and the role of plasmid transmission across different One Health compartments is being described (9). However, methods to compare plasmid similarity are still under development(18,19) and the optimal method to infer plasmid transmission and incorporate tracking of plasmids into IPC outbreak investigations is unknown. This should be a priority for future research.

In contrast to *bla*_NDM-1_ encoded largely on IncHI2/IncHI2A, the *bla*_NDM-5_ gene was encoded by a different IncF and IncR plasmids. Isolates carrying *bla*_NDM-5_ isolates were derived from three species in five hospitals and a community sample and so, in contrast to blaNDM5 (and because of the diverse species and plasmid distribution) there were no strong epidemiologic links with the same species or plasmid. It is possible therefore that transmission of these isolates could represent community acquisition, though without further sampling this must remain a hypothesis. Despite the different plasmid types, nearly identical flanking regions surrounded the *bla*_NDM-5_ including transposable elements, and consistent with a conserved transposable unit circulating in several species (IS*26*-*bla*_NDM-5_-*ble*-*trpF*-*dsbD*-IS*91*). Across England, in hospital settings the *bla*_NDM-5_ has been seen on IncFIA/IncFII multi-replicon plasmids in *E. coli* isolates, as in our study (20)), consistent with national transmission

At the health system level, our study highlights the importance of (and challenges in) tracking antimicrobial resistance or outbreaks across complex healthcare systems. Our hospitals in Liverpool are run by several different hospital trusts, with different data collection systems, different clinicians, managers, and IPC teams. Coordinated action across this system is challenging but our data clearly demonstrate a multi-institution outbreak and highlight the importance of considering the healthcare system as a whole when considering action to tackle AMR. Sharing of data on AMR across organisations is key, and in our case was facilitated by a central microbiology facility.

This work had some limitations, mainly stemming from our use of routinely collected data. Screening for CPEs was non-random, as per UKHSA guidance; the only areas where every patient was swabbed was critical care, in other areas swabbing was based on risk. This will introduce bias, and inflate the number of epidemiologic links in critical care areas compared to other areas. Further information regarding the patient’s stay such as the duration and locations visited within the hospital site was unattainable for some isolates in this study. Admission and ward-based data was not accessible for Hospital 4, Hospital 6 and Hospital 8. This meant we have been unable to identify any potential transmission events within these hospitals for any of our isolates. We lack community surveillance data as CPE screening is targeted at those with recent healthcare exposure so our data do not describe the situation in the community.

## Conclusion

Overall, this work highlights how complex care journeys in modern healthcare systems can combine with the mobility of AMR genes and plasmids to drive spread of AMR. We observed *bla*_NDM_ disseminated by both clonal spread of *K. pneumoniae* ST101, and a successful multispecies IncHI2/IncHI2A plasmid. Patients frequently moving wards and hospitals, including different NHS administrative sections, poses a logistics challenge to find the trade-off between data protection and rapid sharing of relevant information, like diagnostics of highly resistant isolates. To address the novel challenge posed by the AMR crisis it is necessary to consider the problem at the level of the health system, the patient, as well as bacterial species, plasmids and gene cassettes, all of which are essential to resolve to accurately track a complex multi-species multi-hospital outbreak as in our report.

## Supporting information

Supplementary methods, figures and tables except S1

## Data Availability

All metadata used in this study is provided in the supplementary tables, and all raw sequence data is available on SRA/ENA under BioProject PRJEB89345; detailed read accessions are given in Supplementary Table 1.

## Competing interest statement

None to declare

## Author contributions following CRediT taxonomy

Conceptualization: CB, TE, EH. Data curation: CD, DL, JML. Formal analysis: CD, AJF, JML, EH. Funding acquisition: TE, EH. Investigation: CD, DL, VO, LK. Methodology: AJF, FEG, DC, JML, TE, EH. Project administration: JML, TE, EH. Software: CD, JML, EH. Resources: CB, DL, VO, TN, JC, TE. Supervision: AJF, FEG, JML, TE, EH. Validation: DL, VO. Visualization: CD, JML. Writing – original draft: CD, JML, TE, EH. Writing – review & editing: CD, CB, DL, VO, AJF, FEG, DC, JML, TE, EH. All authors read and approved the final version of the manuscript.

## Notes

### Competing Interest Statement

The authors have declared no competing interest.

### Summary of Updates

We have added an author (Lewis Kelly) who contributed to the conjugation experiments; the figures and text were significantly streamlined for ease of interpretation. We improved the definition of strong vs weak links and clarified this in the manuscript, which led to a change in the numbers of these being described.

