## Supplementary methods, figures and tables except S1 for "Multi-species plasmids and *K. pneumoniae* clonal spread driving *bla*_NDM_ outbreak across seven UK healthcare sites"

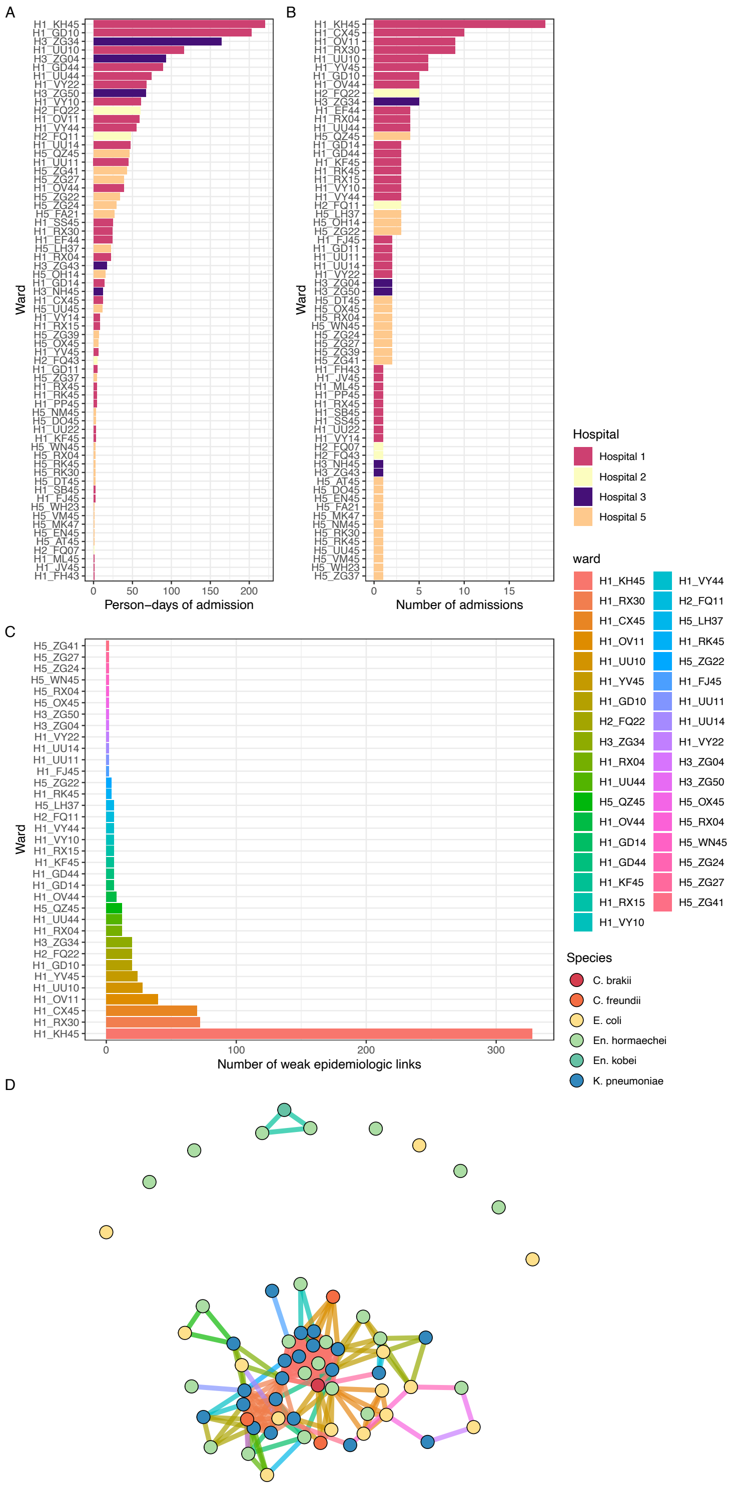


**Supplementary Figure 1:** Person-days of admission **(A)** and number of admissions **(B)** to all the wards in the study; **(C)** number of weak epidemiologic link stratified by ward; **(D)** network plot showing isolates (nodes, coloured by species) linked by edges which represent weak epidemiologic link, coloured by ward where it occurred.

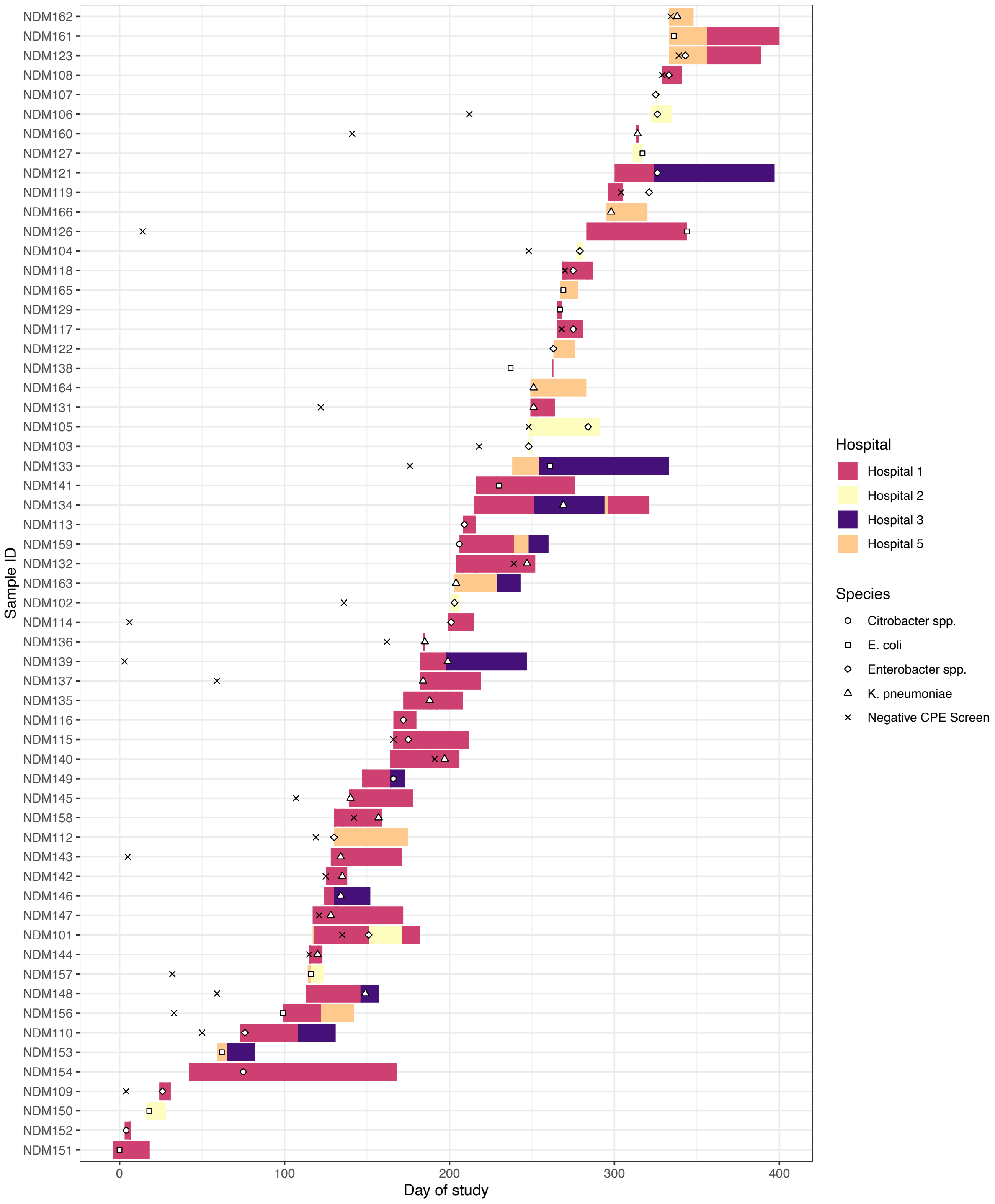


**Supplementary Figure 2:** Overview of sample collection showing duration of hospital admission for all participants (coloured bars indicating which hospital), time of prior negative CPE screening swab (cross) and positive CPE sample (white filled shape).

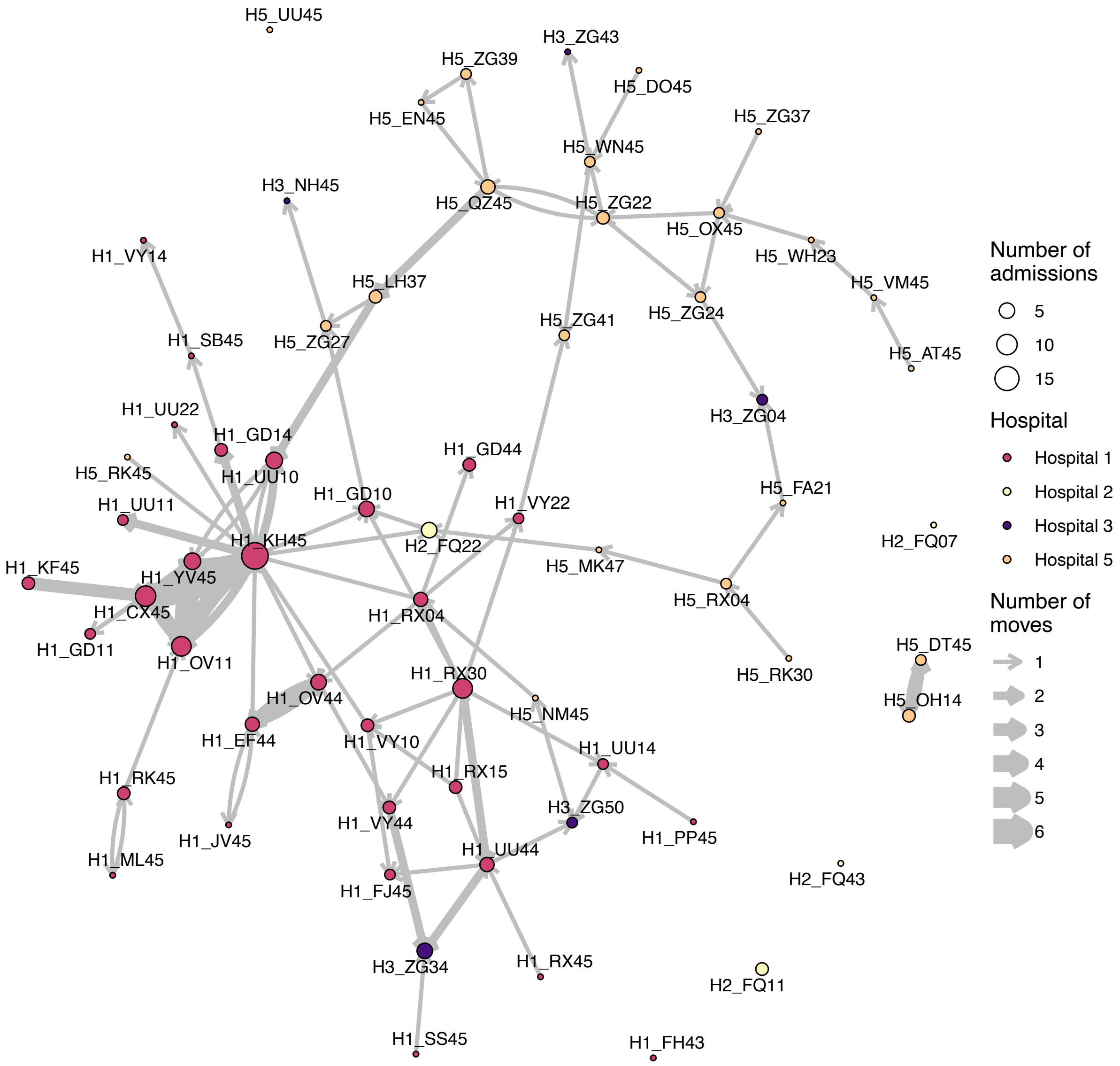


**Supplementary Figure 3: Network plot of all ward moves for patients for whom data is available.** Nodes represent wards, coloured by hospital and size proportional to number of admissions in the dataset. Edges link wards where there has been at least one ward move, with the width of the edge proportional to the number of ward moves.

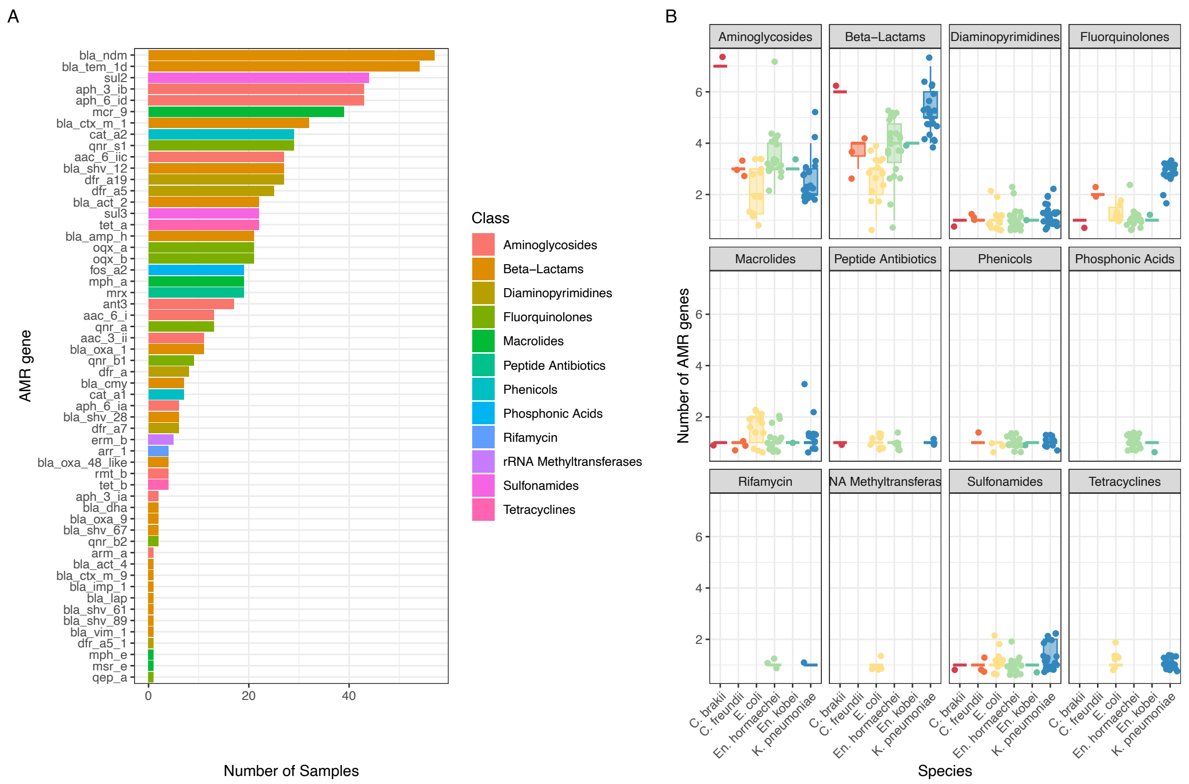


**Supplementary Figure 4: AMR determinants** in our study isolates. **(A)** Number of AMR genes, **(B)** per isolate stratified by species.

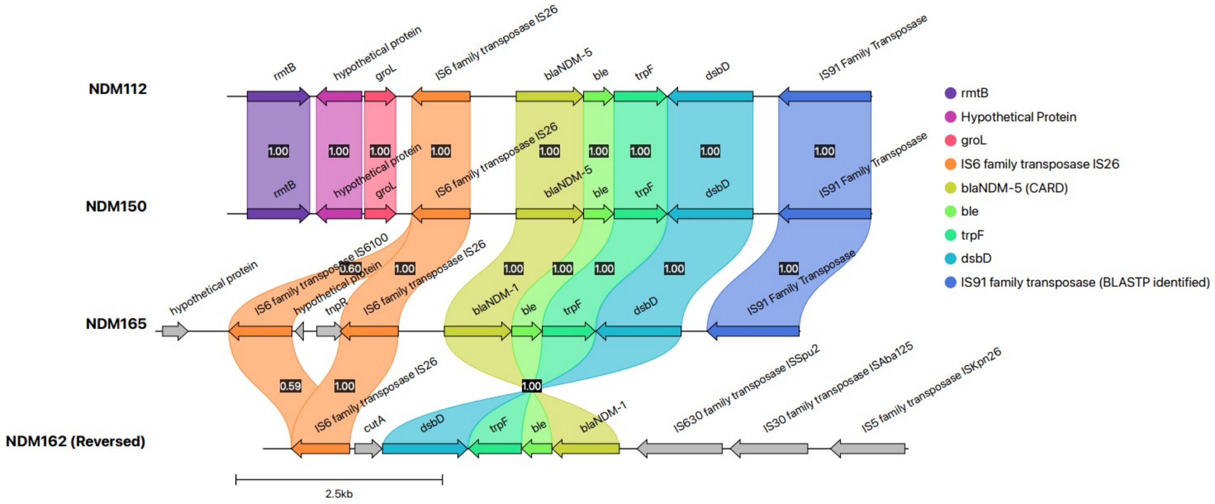


**Supplementary Figure 5:** Genetic context of *bla*$_{\text{NDM-5}}$.

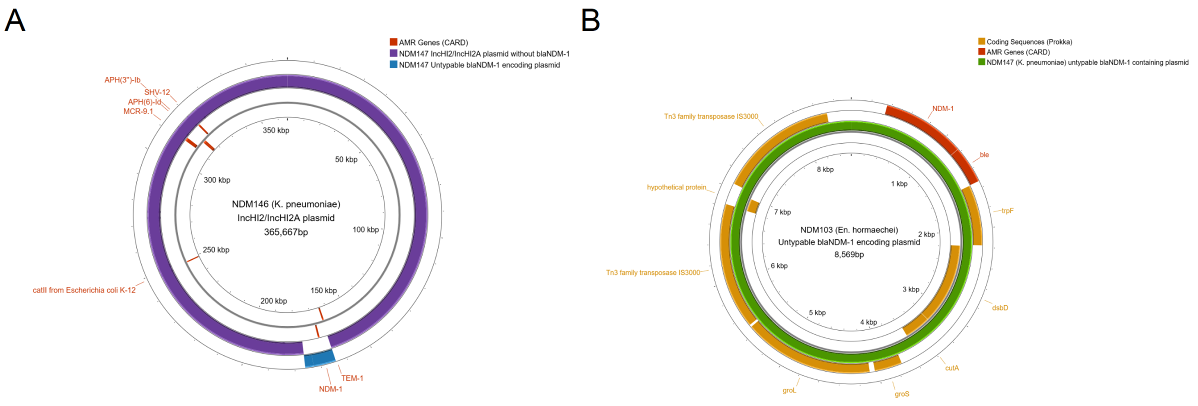


**Supplementary Figure 6: Non-typeable plasmids encoding for blaNDM-1 identified in our collection. (A)** Plasmid sequence from *K. pneumoniae* NDM146, **(B)** from *En. hormachei* NDM103.

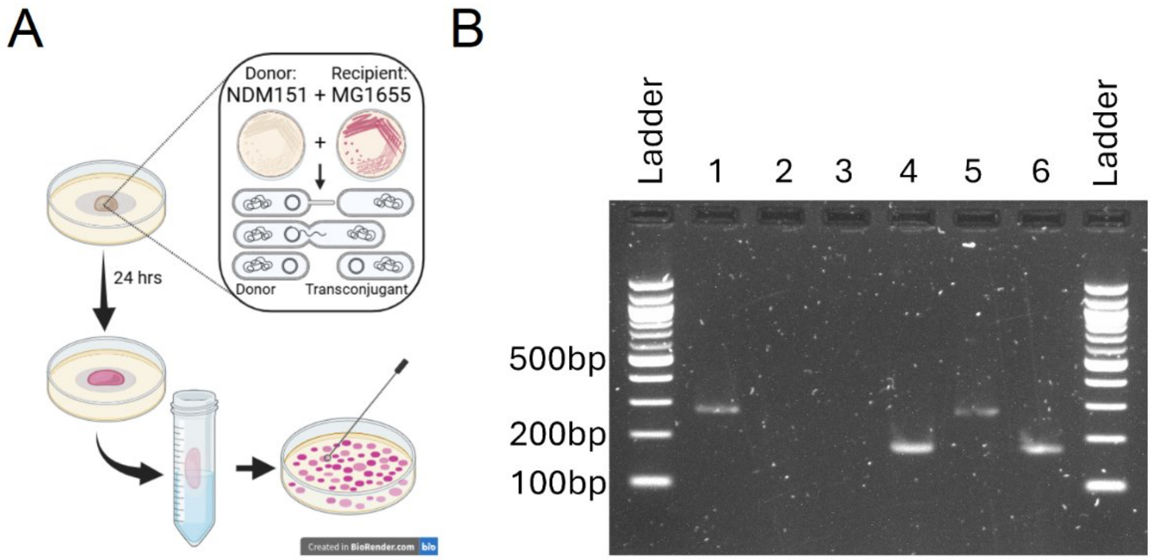


**Supplementary Figure 7: Conjugation assay of plasmid IncHI2/IncHI2A from our study. (A)** Schematic overview of the conjugation experiment between the donor, *E. coli* NDM151, and the recipient, a chloramphenicol resistant *E. coli* K-12 MG1655 harbouring *catA1* gene. **(B)** PCR using *bla*$_{NDM}$ and catA1 primers, column 1 *bla*$_{NDM}$ with NDM151 (donor), column 2 catA1 with NDM151 (donor), column 3 *bla*$_{NDM}$ with MG1655 (recipient), column 4 catA1 with MG1655 (recipient), column 5 *bla*$_{NDM}$ with MG1655 pNDM151_1 (transconjugant), column 6 catA1 with MG1655 pNDM151_1 (transconjugant).

**Supplementary Table 2. Details of hospitals included in this study.**

| Hospital | Services | Unselected Emergency Department | Number of beds |
|---|---|---|---|
| Hospital 1 | Acute hospital, general medical and surgical services | Yes | 857 (source: CQC) |
| Hospital 2 | Specialist oncology | No | 110 (source: clatterbridgec.nhs.uk ) |
| Hospital 3 | Elective surgery, rehabilitation | No | 98 (source: CQC) |
| Hospital 4 | Cardiology, cardiothoracic surgery, respiratory medicine | No | 183 (source: CQC) |
| Hospital 5 | Acute hospital, general medical and surgical services | Yes | 706 (source: CQC) |
| Hospital 6 | Obstetrics, Gynaecology | No | 285 (source: CQC) |
| Hospital 8 | Neurology and Neurosurgery | No | 192 (source: CQC) |

**Supplementary Table 3.** Kleborate prediction outputs.

| Isolate ID | ST | Kleborate virulence score | Yersiniabactin | YbST | Colibactin | CbST | Aerobactin | AbST | Salmochelin | SmST | RmpA DC | RmST | rmpA2 | wzi |
|---|---|---|---|---|---|---|---|---|---|---|---|---|---|---|
| NDM131 | ST101 | 0 | - | 0 | - | 0 | - | 0 | - | 0 | - | 0 | - | - |
| NDM132 | ST101 | 0 | - | 0 | - | 0 | - | 0 | - | 0 | - | 0 | - | - |
| NDM134 | ST101 | 0 | - | 0 | - | 0 | - | 0 | - | 0 | - | 0 | - | - |
| NDM135 | ST101 | 0 | - | 0 | - | 0 | - | 0 | - | 0 | - | 0 | - | - |
| NDM136 | ST101 | 0 | - | 0 | - | 0 | - | 0 | - | 0 | - | 0 | - | - |
| NDM137 | ST101 | 0 | - | 0 | - | 0 | - | 0 | - | 0 | - | 0 | - | - |
| NDM139 | ST101 | 0 | - | 0 | - | 0 | - | 0 | - | 0 | - | 0 | - | - |
| NDM140 | ST101 | 0 | - | 0 | - | 0 | - | 0 | - | 0 | - | 0 | - | - |
| NDM142 | ST101 | 0 | - | 0 | - | 0 | - | 0 | - | 0 | - | 0 | - | - |
| NDM143 | ST3702 | 0 | - | 0 | - | 0 | - | 0 | - | 0 | - | 0 | - | wzi95 |
| NDM144 | ST101 | 0 | - | 0 | - | 0 | - | 0 | - | 0 | - | 0 | - | - |
| NDM145 | ST101 | 0 | - | 0 | - | 0 | - | 0 | - | 0 | - | 0 | - | - |
| NDM146 | ST101 | 0 | - | 0 | - | 0 | - | 0 | - | 0 | - | 0 | - | - |
| NDM147 | ST101 | 0 | - | 0 | - | 0 | - | 0 | - | 0 | - | 0 | - | - |
| NDM148 | ST101 | 0 | - | 0 | - | 0 | - | 0 | - | 0 | - | 0 | - | - |
| NDM158 | ST101 | 0 | - | 0 | - | 0 | - | 0 | - | 0 | - | 0 | - | - |
| NDM160 | ST101 | 0 | - | 0 | - | 0 | - | 0 | - | 0 | - | 0 | - | - |
| NDM162 | ST147 | 1 | ybt 9; ICEKp3 | 173-4LV | - | 0 | - | 0 | - | 0 | - | 0 | - | wzi104 |
| NDM163 | ST101 | 0 | - | 0 | - | 0 | - | 0 | - | 0 | - | 0 | - | - |
| NDM164 | ST147 | 4 | ybt 9; ICEKp3 | 173-2LV | - | 0 | iuc 1 | 63 | - | 0 | - | 0 | rmpA2_6*-55% | wzi64 |
| NDM166 | ST101 | 0 | - | 0 | - | 0 | - | 0 | - | 0 | - | 0 | - | - |

**Supplementary Table 4.** Primers used in the study.

| Target | Forward Primer (5'-3') | Reverse Primer (5'-3') | Reference |
|---|---|---|---|
| ***catA1*** | TTTCGTCTCAGCCAATCCCT | CGACATGGAAGCCATCACAA | (1) |
| ***bla*$_{NDM-1}$** | TGGATCAAGCAGGAGATCAAC | ATTGTCACTGGTGTGGCCG | F: (2) R: This study |

**Supplementary Table 5.** Conjugation frequency data from all biological and technical replicates.

| Experiment repeat | Biological Repeat | Transconjugant colony count | Donor (NDM151) colony count | Transconjugant/donor cell conjugation frequency |
|---|---|---|---|---|
| 1 | 1 | 10 | 10000 | 0.001 |
| 1 | 1 | 90 | 18000 | 0.005 |
| 1 | 2 | 10 | 7000 | 0.001428571 |
| 1 | 2 | 30 | 23000 | 0.001304348 |
| 1 | 3 | 20 | 10000 | 0.002 |
| 1 | 3 | 10 | 27000 | 0.00037037 |
| 2 | 1 | 250 | 66000000 | 3.78788E-06 |
| 2 | 1 | 80 | 40000000 | 0.000002 |
| 2 | 2 | 70 | 60000000 | 1.16667E-06 |
| 2 | 2 | 30 | 44000000 | 6.81818E-07 |
| 2 | 3 | 1 | 69000000 | 1.44928E-08 |
| 2 | 3 | 50 | 63000000 | 7.93651E-07 |

**Supplementary Table 6.** Results for the MIC of meropenem using BMD for the donor, *E. coli* NDM151, the recipient, *E. coli* K-12 MG1655, the *E. coli* transconjugant MG1655 pNDM151_1 and the *E. coli* reference strain ATCC25922.

| **Isolate ID** | **1st Replicate (ug/mL)** | **2nd Replicate (ug/mL)** | **3rd Replicate (ug/mL)** |
|---|---|---|---|
| ATCC25922 | 0.016 | 0.016 | 0.016 |
| K-12 MG1655 | 0.016 | 0.016 | 0.016 |
| NDM151 | 0.125 | 0.06 | 0.125 |
| MG1655 pNDM151_1 | 1 | 1 | 1 |

**Supplementary Table 7.** Predicted features on the hybrid assembly resolved plasmids.

| Isolate ID | Plasmid replicon | Size of plasmid (kb) | AMR genes identified on plasmid |
|---|---|---|---|
| NDM112 | IncR | 46.1 | dfrA12, aadA2, sul1, ble, blaNDM-5, rmtB, blaTEM-1, mphA, APH(3')-Ia |
| NDM150 | IncFIA/IncFII | 92.1 | ermB, mphA, blaTEM-1, rmtB, ble, blaNDM-5, sul1, aadA1, dfrA12 |
| NDM162 | IncFIB(pNDM-Mar)/IncFII(29)/IncHI1B(pNDM-MAR) | 320.3 | ble, blaNDM-5, qnrS1, blaCTX-M-15, dfrA17, aadA5, sul1, mphA |
| NDM165 | IncFIA/IncFIB/Col156 | 137.1 | tetA, tetR, blaCTX-M-15, dfrA17, aadA5, sul1, ble, blaNDM-5, mphA |

### Supporting materials – methods

#### Clinical sample collection and processing

Isolates used in this study came from CPE colonisation screening, and clinical samples where infection was suspected or were collected from the hospital environment as part of an outbreak investigation. Patients were screened using local CPE screening guidelines, following UKHSA recommendations(3). The screening swab was cultured with meropenem selection, and the Gene-Xpert Carba-R assay (Cepheid, United States) was used to identify the CPE gene present in any samples which grew colonies. Clinical samples in which infection was suspected were cultured using standard techniques following UK standards for microbiology investigations (SMI), and speciated with MALDI-TOF mass spectrometer (Bruker, United States) followed by antimicrobial sensitivity testing (AST) using disc diffusion following EUCAST guidelines. For any isolates with a zone size smaller than the EUCAST carbapenemase screening diameter, Gene-Xpert Carba-R assay was used to define presence of carbapenemase genes. Following processing, all isolates were stored on microbank beads at –80C; 64 were retrieved for further analyses.

#### Network analysis

To further explore the movement of patients' where a $bla_{NDM}$ sample was isolated in 2023, the patient's admission and ward location data was extracted from the health record data within the hospital trust, for the admission relating to the isolation of the $bla_{NDM}$ sample and the hospital for any other admissions six months prior to this sample. Where completed, the last negative screening sample was also extracted from patient data. Extraction of this data was only available for Hospital 1, Hospital 2, Hospital 3, and Hospital 5. For isolates that were sampled when the patient was within Hospital 1, Hospital 3, and Hospital 5, the sex and date of birth of the patient was also obtained.

#### Short read sequencing

For short read sequencing, a single colony from each of the isolates was sub-cultured overnight in a Luria-Bertani (LB) broth at 37°C and at 200rpm. DNA was extracted using the DNEasy™ Blood and Tissue Kit (Qiagen, Germany) or the Wizard® gDNA Purification Kit (Promega, United States), and sent to GeneWiz (Azenta Life Sciences, Germany) for sequencing.

#### Long read sequencing

A single colony from each of the 24 isolates selected for long read sequencing (Table S1) was sub-cultured overnight in a LB broth at 37°C and at 200rpm. Two of the isolates (NDM108, NDM159) were resequenced after plating on LB agar (8µg/mL meropenem), as *bla*$_{NDM}$ was not identified from short read sequencing data in these these isolates, and they were able to be grown on 8µg/mL meropenem LB agar plates. After overnight incubation, a single colony was then sub-cultured in LB broth with 8µg/mL of meropenem and incubated overnight at 37°C and at 200rpm. The MasterPure Complete DNA and RNA Purification Kit (LGC Biosearch Technologies, United Kingdom) was used to extract DNA, following the total nucleic acids purification protocol for cell samples. For NDM152, the Wizard DNA Purification Kit (Promega, United States) was used instead due to poor DNA recovery using the MasterPure Complete DNA and RNA Purification Kit (LGC Biosearch Technologies, United Kingdom). DNA was quantified using the Qubit™ Fluorometer (Thermo Fisher Scientific, United States) with the Broad Range Assay Kit (Thermo Fisher Scientific, United States) and the 4150 TapeStation System (Agilent, United States) using the Genomic DNA ScreenTape assay (Agilent, United States). The Native Barcoding Kit 24 V14 (SQK-NBD114.24) (ONT, United Kingdom) and protocol from ONT was used, with all recommended third-party consumables and QC checks. The barcoded library was added to the R10.4.1 Flow Cell on a MinION device (ONT, United Kingdom) before sequencing for 72-hours. Repeat sequencing was completed using the same gDNA and the same ONT kit and reagents, with two isolates per library on a R10.4.1 Flongle (ONT, United Kingdom) for a complete 24-hour sequencing run.

Short read only *de novo* assemblies

FASTQ files were trimmed using the Trimmomatic (v.0.39)(4) paired end option, with the TruSeq3-PE FASTA file of adaptor sequences and recommended parameters. All FASTQ files were processed through FastQC (v.0.11.9) (https://www.bioinformatics.babraham.ac.uk/projects/fastqc/) and collated into MultiQC (v.1.21) (5) for checking. Shovill (v.1.1.0) (https://github.com/tseemann/shovill) was used as the *de novo* short read assembler for reads. QC for assemblies was assessed using CheckM (v.1.2.2) (6) and Busco (v.5.6.1) (7). Isolates that had a value greater than 5% for contamination in CheckM was inspected manually using the Artemis Comparison Tool (V.13.0.16) (8) using other isolates within the collection as references. GAMBIT v.1.0.0 (9) was used to confirm the species of the isolates, by comparing them to a NCBI derived database produced by GAMBIT (downloaded 29$^{th}$ April 2024).

Long read *de novo* assemblies

All long reads were basecalled using the guppy super accurate model option on a GPU (v.6.4.6), and demultiplexed using a CPU (v.6.5.7). The Hybracter (v.0.7.3) (10) assembly pipeline was used to assemble all long read sequences, using Flye (v.2.9.3) (11) as the assembler, and polish the assemblies with the long and short read sequences described above. Due to low coverage from long reads in eight isolates, Unicycler (v.0.5.0 or v.0.5.1) (12) was used as an assembler with the same filtered and trimmed long and short reads, produced from the Hybracter pipeline, as input. Assemblies were polished with Medaka (v.1.11.3) (https://github.com/nanoporetech/medaka), Polypolish (v.0.6.0) (13) and Pypolca (v.0.3.1) (14) with the --careful parameter. All draft assemblies were checked using CheckM (v.1.2.2) (6); continuity of the presence of AMR genes and plasmid replicons were checked between the short read and long read derived assemblies using StarAMR (v0.10.0) (15).

### Genomic analysis

All final assemblies were annotated using Prokka (v.1.14.6) (16). Mash (v.2.2.2) (17) was used to produce a file of the estimated genome distances between all isolates for use in hierarchical clustering within heatmaps. MLST were called using ariba (v.2.14.6) (18) with the PubMLST typing schemes for *Citrobacter freundii*, *Enterobacter cloacae* complex (ECC), *E. coli* (Achtman's seven-gene scheme), and *K. pneumoniae* (all downloaded 9$^{th}$ May 2024).

The Panaroo (v.1.5.0) (19) pan-genome pipeline was used to calculate the pan genome for each of the assemblies using the '--clean-mode strict' and '-a core' options. From the core gene alignment produced, SNP-sites (v2.5.1) (20) was used to extract only SNPs within the alignment and the number of constant sites. These files were passed to IQ-TREE (v.2.4.0) (21) to produce a best fit maximum likelihood tree file for the genus using a general time reversible model for all trees. These trees were then visualised using iTOL (v.7.2) (22).

A SNP-based phylogram of ST101 *K. pneumoniae* isolates was generated using Snippy (v.4.6.0) (https://github.com/tseemann/snippy), with the chromosome from the final long read assisted genome assembly of NDM144 as the reference, and the trimmed short reads for the rest of the ST101 isolates. Pairsnp (v.0.3.1) (https://github.com/gtonkinhill/pairsnp) calculated the pairwise SNP distances of each ST101 isolate. The core alignment produced by Snippy was then cleaned using snippy-clean_full_aln. Gubbins (v.3.3.1) (23) was run using the GTRGAMMA model. SNP-sites (v.2.5.1) was then used to produce a FASTA file of only SNPs between the isolates after recombination were removed. RAxML-NG (v.1.2.2) (24) was used to produce a general time reversible model, maximum likelihood tree. Pyjar (https://github.com/simonrharris/pyjar), as implemented for python (v.3.7), was used to produce a joint ancestral reconstruction tree from the best maximum likelihood tree outputted from RAxML-NG. Based on community standards, a sub-lineage threshold of 5 SNPs and a threshold of 3 ancestral nodes to the root was selected for rPinecone (v.0.1.0) (25) . The SNP scaled tree, generated using the pyjar derived tree file and alignment, was then visualised in

R (v.4.3.1) using the R packages ggtree (v.3.10.0) (26), ggplot2 (v.3.5.0) (27) , and patchwork (v.1.3.0) .

Kleborate (v.2.3.2) (28) output, with the implemented SRST2 (29), was used to predict virulence genes for each *K. pneumoniae* isolate. The CARD and SRST2 databases (download dates 14th February 2024 and 3rd May 2024 respectively), were used when running ariba (v.2.14.6) to look for AMR genes in the isolates and the ResFinder database (downloaded 3rd May 2024) was used alongside the other databases to check for consistency in the $bla_{NDM}$ variant identified. From the ariba AMR results using the SRST2 database, any results for the *catBx* gene were excluded from data analysis (1). To assess the presence of plasmid replicons, the PlasmidFinder database (30) (download 24th April 2024) was used with ariba. Ariba results for both AMR genes and plasmid replicons were visualised in R using the pheatmap package (v.1.0.12).

Abricate (v1.0.1) (https://github.com/tseemann/abricate) was used with both the CARD and ARG-ANNOT databases (download June 2024) to confirm the $bla_{NDM}$ variant and the contig it was on. StarAMR (v0.10.0) was used to confirm the plasmid replicons present and the plasmid replicons of the plasmid of the $bla_{NDM}$ contig. Bandage (v.0.8.1) (31) was used to export the nucleotide sequence of the plasmid of interest to Proksee (32) to visualise in a circularised figure. Within Proksee, the CARD Resistance Gene Identifer tool (tool v.1.2.1) was used to annotate the plasmid figure with AMR genes; these genes were manually checked for consistency with Abricate results. The BLAST tool (tool v.1.3.1) was used to compare and visualise the plasmid sequences of different isolates.

Minimap2 (v.2.24) (33) and samtools (v.1.19.1) (34) was used to map reads back to an assembly and process the alignment to check to ensure reads were represented accurately in the assembly; Tablet (v.1.21.02.08) (35) was used to visualise the bam file against the assembly. To compare regions of certain isolates of interest, Artemis (v.16.0.17) was used on the annotated gff file to select a region upstream and downstream from the gene of interest. This region was then re-annotated with Prokka (v.1.14.6), before being compared and visualised with Clinker (v.0.0.29) (36).

### Conjugation of IncHI2/IncHI2A

To test the ability for conjugation of a $bla_{NDM-1}$ IncHI2/IncHI2A plasmid filter mating was performed using NDM151 (*E. coli*) as the plasmid donor with a chloramphenicol resistant MG1655 (*E. coli*) strain harbouring a *catA1* gene on a small non-mobilisable plasmid that also expressed a red/pink colour (eforCP gene) as the recipient. Each conjugation assay was performed in duplicate with three biological replicates per repeat. In short, donor and recipient cells were grown overnight in LB broth with meropenem and chloramphenicol selection, respectively, at 37 °C with 220 rmp shacking. Overnight cultures were washed once in prewarmed LB broth medium to remove residual antibiotics. The donor was diluted 1:20 in fresh LB and incubated until early to mid-exponential phase was reached($OD_{600}$ = 0.3-0.6). Donor and recipients were transferred into a microcentrifuge tube, spun down at max speed for 2 min, and cell pellets resuspended in 50 µl M9 salts solution. Donor and recipients were then mixed in equal volume and transferred to a 0.22µM nitrocellulose membrane filters (Merek, Germany) that had been placed on a LB agar plate and incubated for 24 hours incubation at 30°C. Then, the filters was placed on a microcentrifuge tube and cells were suspended in 1 mL cold M9 salts by vertexing. This was then serially diluted in cold M9 salts (up to $10^6$) and 10 µL spots plated in duplicates on LB meropenem + chloramphenicol plates to select for transconjugants and meropenem plates to select for donors. Transconjugants are also able to grow on meropenem plates but are several orders of magnitudes lower than donor and should appear as red/pink colonies, we thus assumed all colonies are higher dilution to be donors. These plates were incubated at 30°C for a 24 h before counting donor and transconjugants. The conjugation frequency was then calucated as the ratio of transconjugants/donor. To confirm transconjugants, alongside the visual colour and being resistant to meropenem and chloramphenicol, colony PCRs were done using the DreamTaq PCR Master Mix (2X) (Thermo Fisher Scientific, United States); primers used in these PCRs and their source are described in Supplementary Table 4. Results were visualised on a 1% agarose gel. Transconjugants from the successful *E. coli* donor (NDM151) to the MG1655 *E. coli* strain recipient conjugation expressed the pink colour of the recipient and colony PCR confirmed that the transconjugant MG1655 pNDM151_1 contained both the $bla_{\text{NDM-1}}$ gene from the donor and the catA1 gene from the recipient.

Meropenem susceptibility testing

The minimum inhibitory concentration (MIC) of meropenem (ApexBio, USA) was identified for the donor (NDM151, *E. coli*), recipient (MG1655, *E. coli*) and a transconjugant (MG1655 pNDM151_1), with three technical replicates performed. Due to concerns regarding the loss of blaNDM before MIC testing, both NDM151 and MG1655 pNDM151_1 were attempted to be grown in Muller Hinton agar supplemented with 4ug/mL of meropenem. This was successful for MG1655 pNDM151_1 but unsuccessful for NDM151. Susceptibility testing was completed using the MIC broth microdilution method, by passaging single colony from a Muller Hinton agar plate in 10mL Millipore NutriSelect® cation-adjusted Muller Hinton broth (Merek, Germany) for 8-18 hours, 200rpm, at 37C. This was then diluted to an OD600 of 0,001 (corresponding to 10 5 cfu/mL) in cation-adjusted Muller Hinton broth. To prepare MIC plates, 96 round bottom well microtiter plates had meropenem serially diluted across an eighteen-fold dilution range in cation-adjusted Muller Hinton broth in triplicate wells per drug concentration. Equal volumes of E. coli inoculum were added to wells containing serial diluted meropenem, including drug free wells as positive growth controls. Inoculum free cation-adjusted Muller Hinton broth wells were also included as media contamination wells.

Additional information on untypeable plasmids

Compared in BLASTn using the megablast algorithm with a 100% identity and 100% query cover. Both elements were shown in the circular element comparison. These circular elements were not identified to contain any phages, using PHASTER.

In a search against the core nucleotide BLAST database of assemblies, the untypable small element of NDM103 showed 99.4% identify and 100% query cover to a larger 304.6 kbp *bla*$_{NDM-1}$ encoding *K. pneumoniae* plasmid pKJNM8C2.1 (accession CP030858.1) collected from a neonate in India in 2017, and the untypable small element of NDM147 had 100% identity and 100% query cover to the larger 62.7kbp *bla*$_{NDM-1}$ encoding *E. cloacae* plasmid pNDM1-CBG collected from a sputum sample in China in 2017 (accession CP046118.1). Further investigation of these sequences using the BLASTX program with the non-redundant protein sequences (nr) database to identify protein-coding sequences showed hits in both sequences to GroEL (COG0459), TrpF (COG0135), GroES (COG0234), *bla*$_{NDM-1}$ (cd16300), GloA (COG0346), CutA (COG1324), DsbC (pfam11412). The Tn3-like element IS3000 family transposase (NF033527) and IS30 family transposase (COG2826) were also identified in both.
